# Long Covid and the role of physical activity: a qualitative study

**DOI:** 10.1101/2020.12.03.20243345

**Authors:** Helen Humphreys, Laura Kilby, Nik Kudiersky, Robert Copeland

## Abstract

**Objectives:** To explore the lived experience of Long Covid with particular focus on the role of physical activity

**Design:** Qualitative study using semi-structured interviews

**Participants:** 18 people living with Long Covid (9 male, 9 female; aged between 18-74; 10 White British, 3 White Other, 3 Asian, 1 Black, 1 mixed ethnicity) recruited via a UK-based research interest database for people with Long Covid

**Setting:** Telephone interviews with 17 participants living in the UK and 1 participant living in the US

**Results:** Four themes were generated. Theme one highlights the physical and social isolation experienced by people with Long Covid, compounded by a lack of support and advice from medical professionals. Theme two describes how participants sought information and validation through online sources and communities. Theme three captures the challenges associated with managing physical and cognitive effects of Long Covid including fatigue and ‘brain fog’ whilst trying to resume and maintain activities of daily living and other forms of exercise. Theme four illustrates the battle with self-concept to accept reduced function (even temporarily) and the fear of permanent reduction in physical and cognitive ability.

**Conclusions:** This study provides insight into the challenges of managing physical activity alongside the extended symptoms associated with Long Covid. Findings highlight the need for greater consensus around physical activity-related advice for people with Long Covid and improved support to resume activities considered important for wellbeing.

**Article Summary:** 

**Strengths and limitations of this study:**

- To our knowledge, this paper is the first to explore the role of physical activity in the lived experience of Long Covid using a qualitative approach
- The study design enabled in-depth inquiry of lived experiences in a diverse sample
- Inductive thematic analysis ensured descriptions and interpretations of the lived experience were tested and found to be grounded in the data
- Participants were recruited from members of a Long Covid research interest database who registered via an on-line form, meaning study findings might not capture the views of digitally excluded populations

**Funding statement:** This work was supported by Sheffield Hallam University.

**Competing interests:** All authors have completed the ICMJE uniform disclosure form at www.icmje.org/coi_disclosure.pdf and declare: no support from any organization for the submitted work; no financial relationships with any organizations that might have an interest in the submitted work in the previous three years; no other relationships or activities that could appear to have influenced the submitted work.

## INTRODUCTION

To date, more than 30 million cases and 1 million deaths of COVID-19 have been reported worldwide (1). The medical and research community has focused on understanding COVID-19 pathophysiology and supporting treatment of acute cases of COVID-19 (2). Whilst the majority of people infected recover, a significant proportion experience ‘Long Covid’, showing severe symptoms for weeks and even months post-infection, irrespective of age and in the absence of underlying health conditions. Long Covid appears to be a multi-system disease associated with a complex array of respiratory, neurological, cardiovascular, gastrointestinal, musculoskeletal, rheumatological, dermatological and immunological symptoms ranging in severity, frequency and duration (3-6). Preliminary findings from magnetic resonance imaging investigations also show that ∼70% of ‘low-risk’ individuals testing positive for COVID-19 present signs of impairment in one or more organs four months after symptom onset (7). Research highlights common characteristics of Long Covid including severe fatigue and impaired physical and cognitive function, inhibiting activities of daily living (3,8,9). Engaging in physical activity (PA) has been reported to trigger the onset of acute symptoms (e.g. rapid heartbeat and shortness of breath) and post exertional malaise, (3). One hypothesis is that persistent symptoms are caused by organ dysfunction induced by the virus, potentially compounded by deconditioning of physical fitness as a result of sedentary behaviour (10-12). Physical exertion exacerbates fatigue and higher likelihood of prolonged sedentary periods (3,8) creating a negative cycle.

The response to Long Covid is accelerating, reflected in a dynamic themed review by the National Institute of Health Research (NIHR) on ‘Living with Long Covid’ (9) and NHS England plans to establish of ‘post-COVID syndrome’ clinics (13). To date, there has been no formal research exploring the role of PA in the management and rehabilitation of Long Covid. This study explores the lived experience of people with Long Covid including the role of PA -to inform the design and implementation of rehabilitation support.

## METHODS

### Participants and recruitment

Interviews were conducted with English-speaking adults aged >18years who self-identified as recovering from COVID-19, were not hospitalised (did not receive in-patient treatment) and had experienced a recovery period lasting 3 weeks or more, a timeframe consistent with definitions of ‘post-acute COVID-19’ at the time of study initiation (14). Ethical approval was granted by Sheffield Hallam University. Participants were recruited from a COVID-19 research interest database (the RICOVR database) established by the Advanced Wellbeing Research Centre (AWRC) at Sheffield Hallam University (15). At the time of the study, 2023 people were registered on the AWRC database. In line with current guidance, a positive COVID-19 test was not a prerequisite for participation (14, 16).

Purposive selection was used to ensure that the sample reflected a range of ages, genders and ethnicities. Database members were stratified by age group, gender and ethnicity, then selected chronologically according to the date that they registered with the RICOVR database. Two rounds of invites were sent by email; if invitees did not respond after two weeks or decided not to participate, we sent a new invite to the next person registered on the database (demographically matched). A total of 35 people were invited to participate; 21 responded to indicate interest. Respondents were provided with full details about the research and invited to an informal telephone discussion with the interviewer (HH) to discuss the research aims and procedures. 18 people took up this opportunity; all subsequently decided to proceed. Written consent was collected from all participants prior to interview. 17 participants were residing in the UK one was living in the US. After interviewing these 18 participants, the research team were satisfied that no new themes were being identified and recruitment ceased.

### Patient and public involvement

During study design, the AWRC Public Involvement in Research Group (17) reviewed study aims and all materials. The group provided feedback to refine documents including clarification of language in the participant information sheet, rewording of interview questions and the addition of information about support for carers in the post-interview support document.

### Interview procedures

A semi-structured interview guide was developed to elicit participants’ stories about their lived experience of Long Covid and the role of PA within that experience. Open questions explored four broad topic areas: (i) illness and recovery trajectory, (ii) sources of support (iii) experiences of PA (iv) future priorities and concerns. All interviews were conducted during September and October 2020 via telephone with the exception of 2 interviews carried out using Zoom video conferencing to suit participants’ needs. All interviews were conducted by HH, an experienced qualitative researcher in public health and exercise psychology.

Interviews were audio recorded and limited to a maximum of 45 minutes to limit any cognitive burden for participants. Participants were not reimbursed but were signposted to information detailing sources of support within and beyond the University should any distress have been caused by the interview.

### Data analysis

All recordings were transcribed verbatim by a professional transcription service. Transcripts were sent to participants for review; one participant responded with clarifications which were included in our analysis. Reflexive thematic analysis with inductive, semantic coding (18) was used to interpret the data. Consistent with recommendations, we did not set out to achieve inter-coder reliability (19). Instead, multiple researchers coded the transcripts to encourage reflexivity and ensure our analysis considered different possible interpretations. Two researchers (HH and LK, both with postgraduate psychology qualifications, training and experience in qualitative interviewing) reviewed 50% of the transcripts each. HH and LK independently developed preliminary coding frameworks presenting initial themes, which they compared and refined with input from a third researcher (NK) who had read a cross section of the transcripts. Following discussion with a fourth researcher (RC) HH, LK and NK returned to the transcripts to sense-check candidate themes and ensure that they offered an appropriate representation of the data, at which point themes were defined and named. Final themes are presented below along with illustrative participant quotes.

## RESULTS

### Participants

Table 1 displays sample characteristics for the 18 people who participated.

**Table 1:** Participant Characteristics.

| ID | Sex | Age group | Ethnic group | Place of residence |
| --- | --- | --- | --- | --- |
| 1 | Male | 55-64 | White - Irish | UK |
| 2 | Female | 45-54 | Asian or Asian British - Indian | UK |
| 3 | Male | 35-44 | White - British | UK |
| 4 | Male | 18-24 | Black or Black British - Caribbean | UK |
| 5 | Male | 45-54 | White - British | UK |
| 6 | Male | 65-74 | White - British | US |
| 7 | Female | 25-34 | Mixed - White & Asian | UK |
| 8 | Female | 65-74 | White - British | UK |
| 9 | Male | 35-44 | Asian or Asian British - Indian | UK |
| 10 | Male | 65-74 | White - British | UK |
| 11 | Female | 18-24 | White - British | UK |
| 12 | Male | 18-24 | White - British | UK |
| 13 | Female | 65-74 | White - British | UK |
| 14 | Female | 55-64 | White - British | UK |
| 15 | Male | 45-54 | White - Other | UK |
| 16 | Female | 35-44 | White - Other | UK |
| 17 | Female | 35-44 | White - British | UK |
| 18 | Female | 45-54 | Asian or Asian British - Indian | UK |

### Physical, social and medical isolation

All participants described a profoundly isolating experience. Most participants reported difficulty accessing healthcare services during the initial phase of their illness. Many felt their symptoms were not serious enough to warrant emergency care, yet access to GP services was denied, delayed or limited:

> *“when you phoned your GP up they just referred you back to 111. So I was in this cycle of not being able to get any help, so all these symptoms were coming out and I just didn’t know who to turn to really” (IV10)*

Participants described significant debilitation, with their physical function drastically reduced and in most cases, at least several weeks of being virtually housebound. Basic activities of daily living including self-care and housework became challenging or impossible:

> *“The slightest thing was an effort in a way I’ve never ever conceived before, it’s the most fatigued I have ever been*… *things like changing my bedding, I did in stages like one pillow case and then later in the day I’d do another pillow case, it was that sort of level of difficulty with day-to-day tasks*.*” (IV2)*

Along with national lockdown restrictions, this constituted a physical isolation compounded by limited public health messaging and media coverage about Long Covid that created an additional sense of social isolation. Public health messages were seen as portraying the COVID experience in binary terms: either requiring hospitalisation or being ‘mild’ enough to recover at home within a short time frame (2 weeks). Meanwhile, media outlets primarily reported mortality, hospitalisation and new case statistics. Neither narrative matched the experience of our participants:

> *“Unless I’m wrong, I don’t think the government have said anything. They don’t put it in their statistics. So they talk about death rate, they talk about hospital cases and they talk about new cases, but they don’t talk about people who are still struggling months on*.*” (IV7)*

> *“For quite a long time while I was lying at home floored by this; they were just saying that younger people should be fine and that it’s the older generation that we need to protect and it was just, I felt, kind of selfish and a bit like well that’s just wrong*…*” (IV11)*

The sense of isolation was most acutely felt through the lack of answers and support forthcoming from frontline healthcare professionals. All participants reported feeling variously let down by the level of practical intervention offered to them (e.g. direct treatment; tests and access to results), limited medical knowledge and awareness of Long Covid. Most participants reported that interactions with healthcare professionals fell well below meeting their psychological support needs (e.g. feeling believed, space to talk through their worries, advice about the likely disease path):

> *“They [doctors] didn’t know how to handle the information. The guidance wasn’t there for them. They didn’t know who to refer to. They didn’t know if they could refer*… *But I did not feel heard. And I think as a patient I felt extremely lost from what should have been a service that could signpost and support and recognise distress and uncertainty, there was very little acknowledgement of that*.*” (IV9)*

Others conveyed that although medical advice was limited, they appreciated being listened to and believed. Although early medical interactions had been disappointing, there was recognition that medical knowledge was catching up. Indeed, the penultimate participant we interviewed had been formally diagnosed by her GP as having Long Covid, reflecting an emerging understanding amongst clinicians.

### Seeking validation and answers

The lack of understanding and explanation from trusted support sources led all participants to seek information and validation online. Social media forums provided a community which normalised the experience and suggested coping strategies:

> *“People post videos, talks, articles, and that has been my main source of information… So rather than sitting round worrying, I’ve known that this is a problem, it’s not just me being a bit anxious or a bit of a hypochondriac, it’s normal if you’ve had this virus, and that’s been brilliant” (IV12)*

In addition, most participants adopted a researcher role, reading scientific articles and health resources to better understand what was happening to them:

> *“And then obviously I’ve been reading a lot of evidence papers as well. So I’ve been looking up stuff like that as well and trying to get the research from that side of things and trying to form my own opinion and diagnosing myself” (IV18)*

Professional advice often arrived after information had already been accessed online:

> *“They [physiotherapist] offered loads of advice just about pacing really. But I think at that point because of all the communities that have sprung up everywhere online people had already been sharing this information*.*” (IV3)*

Whilst the online research and social media communities were broadly deemed as supportive, they could also lead to anxiety, by accentuating negative experiences and creating doubt about longer term prognoses:

> *“So I initially found it very useful, because I didn’t feel like I was making it up, I didn’t feel like I’m on my own completely here. But now I’ve backed off from those groups, because there are some really horrid stories. And when I’m mentally low I don’t need to hear how other people are really struggling and have it as well. And then also some people are a few months ahead of me in the support groups. And I want to have a little bit of hope that I’ll get better. But if I see people still struggling at seven/eight months and two months ahead of me, they’re worse off, it just doesn’t help me*.*” (IV7)*

### Learning how to balance symptoms and activity

All participants described physical and/or cognitive fatigue which came in cycles or episodes. This manifested as physical lethargy, a lack of coordination and/or brain fog resulting in significant debilitation and often confinement to bed:

> *“I felt from the very beginning that I was in a cyclic washing machine if you like, because I would say on a two week basis I was seeing the symptoms recycling*.*” (IV1)*

> *“I’m used to feeling tired, feeling fatigued, but this is on another level. I’ve gone from being able to go out and run; at times I couldn’t cross the room. I struggled to get upstairs*… *I described it as wearing a suit of armour*…*and on top of that I get this lack of coordination. I can’t grip things. I can’t manipulate things with my hands. It’s like trying to do things wearing ski gloves… And then, on top of that, it is this brain fog*. *It’s like*… *in my younger days, when I was drunk*…*you have to focus and you have to do things very slowly and carefully*.*” (IV5)*

Consequently, participants described losing the freedom to engage with routine activities. Any physical or cognitive activity could result in the onset of severe fatigue, resembling post-exertional malaise. In addition, PA often triggered acute symptoms including heart palpitations, breathlessness, joint and muscle pain:

> *“So if I do something physical I suffer. If I walk I suffer in my legs. If I do something with my hands I suffer with my hands. If I start to think too much I then get a foggy head. If I type an email on the computer and it goes on too long, I then can’t think enough to shut the computer down*.*” (IV5)*

Participants differed in their attitudes to these relapses. Some considered them worthwhile, either because with each relapse followed a small improvement in baseline function, or it was considered a price worth paying for the sense of normality, control and positive affect that the activity provided:

> *“So as much as I’m enjoying [walking the dog], it has the knock on effect. But that is getting less and less, so the more I’m doing the better I’m feeling afterwards. I think [relapses are] all part of it, just got to get on with it and push myself a little bit harder and then hopefully I’ll get better quicker. It doesn’t put me off*.*” (IV17)*

Others believed it was not worth the risk and feared the potential of long-term damage that could be caused:

> *“…I’ve always been one of those people that things well, you know, you push through it…*.*But this you just can’t. And this is something that I’m becoming more afraid of that I think maybe I need to properly back off from as much daily activity as I can to recover from this because I’m scared that I will eventually end up as probably a 50% to a 60% of what I was previously, permanently, or for a longer term*.*” (IV5)*

Medical advice regarding physical activity was sparse. Most participants had experimented with graded approaches to exercise and activity, using resources found online, although for many this advice had been confusing. Whilst navigating conflicting sources of advice, other challenges included difficulties establishing a safe, consistent baseline for activity amidst daily unpredictability in symptoms:

> *“There’s obviously people that have had… different types of viruses, and they’re all claiming that doing exercise and whatnot is harmful for your recovery*.*” (IV04)*

> *“Everything that you read is pace yourself, pace yourself. But that’s really hard to do, because until you’ve overdone it you don’t know how much you can do without overdoing it, if you see what I mean, so learning by default. Which isn’t the best way, but I guess what’s enough for me might not be enough for somebody else*.*” (IV08)*

### Adapting to an altered life

There is a clear sense that this illness is experienced as life altering. Many participants described a loss of ‘self’ and/or narrated a substantial impact on their identity. Some participants made sense of this as a ‘pre’ and ‘post’ COVID life, others described it as a journey, and all were struggling with the notion that this changed self may or may not become permanent:

> *“My biggest concern is that nobody knows the prognosis. I’m hearing some people getting better and some people aren’t and my biggest worry throughout all of this is if this is it, if this is going to be permanent… But it’s just that not knowing and being in limbo for such a long time and for the foreseeable that I find the most difficult*.*” (IV16)*

The prospect of permanent disablement was distressing for all participants, but whereas older participants drew on life events such as previous illness to make sense of their current experience, we found that younger participants (e.g. those 18-24) particularly struggled with their incapacitation, coupled with the loss of their usual face-to-face social networks and coping mechanisms:

> *“I just want my life back, it’s getting a bit tedious. I see myself just becoming a burden, I don’t want to live my life like that. I don’t want to feel like a burden to my mum, I just want to go back to life” (IV4)*

The majority of participants had been unable to resume activities that were previously central to their core identity (e.g. a parent, an employee, an active person). Anything that provided a sense of normality helped to refute the idea that this new identity was permanent (a prognosis that was both feared and resisted). Participants for whom PA was a core feature of their self-concept prior to contracting COVID-19 referred to this as a strong motivator in their desire to improve functional capacity and return to pre-Covid PA behaviour. Awareness that they had previously been able to achieve high levels of fitness fostered a belief that at least some return to fitness might be possible:

> *“I just wanted to go in the garden*… *I wanted to be normal. I think normality was a massive thing in my head*.*”(IV14)*

> *“I’ve been a runner and then a cyclist for many years so I had the intent of getting back in the walking. And then as soon as I could I got back in the cycling a little bit”. (IV06)*

Some participants had reached a point of ‘reluctant acceptance’, not necessarily arrived at peacefully but through exasperation and for some, a degree of self-defeat. Participants described needing to give themselves permission (or seeking it from others) to rest and adjust their energy expenditure and lifestyle accordingly:

> *“I think I’ve just got to the point where I’ve accepted my new norm, so I’ve just been told rest and give it time*.*”(IV17)*

> *“I tend to be fine, I’ll just go out and exercise and recover and nothing’s really held me back whereas this has humbled me and made me realise I need to be more careful. So I guess there’s a bit of self-learning going on there. But I would rather be ignorant and healthy but that’s not really an option*.*”(IV15)*

Participants’ lives had been so disrupted that many had experienced a re-examination and shift in priorities:

> *“I feel I have learnt a lot about my own resilience…And I feel there’s an opportunity for change. I might reduce my hours going forward. It’s difficult but I might try and balance my work-life balance a bit more and pace myself*.*”(IV9)*

## DISCUSSION

### Addressing the impact of Long Covid

Participants described isolating experiences exacerbated by fragmented and largely unsupportive medical care which echo previous reports (20). Whilst participants acknowledged the difficulties associated with diagnosing and treating a novel and undetermined syndrome, early care experiences had negatively impacted many participants’ physical and emotional health. This reflects research indicating that survivors of COVID-19 could be at increased risk of adverse mental health including anxiety (21). Our findings substantiate the need for holistic support addressing the physical and psychological impacts of Long Covid, reflected in guidance for the establishment of “post-COVID syndrome assessment clinics” (13).

### Resuming and maintaining ‘normal’ activity

Activities of daily living (e.g. housework, gardening) and outdoor activity were referred to as crucial links to normality, and vital for mental health. Our findings indicate that people experiencing Long Covid need better support to manage their symptoms, especially fatigue, whilst also helping them safely pursue the benefits of PA that were so badly desired. This might include support to establish a baseline and upper threshold for activity which takes into account the apparent relapse-recovery cycle common to our participants’ experience. People with Long Covid need to feel competent and confident to apply principles of pacing and many will require monitoring to provide reassurance about the safety of PA whilst experiencing other symptoms like rapid heartbeat or shortness of breath. Given the complexity of the recovery process – particularly in terms of PA - the direct involvement of people with Long Covid in the design of services to support recovery appears critical.

Our findings reflect concerns regarding the potential risk of long-term damage associated with post-exertion malaise and PA. Participants differed in their attitudes towards relapse, some believing they were constructive to recovery whilst others feared danger to long-term health. This paradoxical role of PA in relation to relapse and recovery reflects previous qualitative studies involving people with physically limiting conditions such as multiple sclerosis (22). Parallels have been drawn between Long Covid and myalgic encephalomyelitis (ME) and/or chronic fatigue syndrome (CFS) (23). Recently, NICE withdrew a recommendation to prescribe graded exercise therapy for patients with ME/CFS following concerns it could cause harm to some patients (24). It is imperative to establish consensus, adding to what is already known (14) regarding PA-related advice specifically for people with Long Covid, including the identification of individual phenotypes for whom PA might or might not add value to their recovery.

### Access to information and the role of the internet

Isolation and a lack of support left our participants with no choice but to self-manage and self-organise. The internet offered a crucial tool for accessing support, validation and information about how to manage the Long Covid experience. This information was disseminated much faster online than it could be filtered through to frontline GPs. Previous research suggests that suggests that online support communities can readily address the support needs of people with long-term conditions that are currently unmet offline (25). The ability of online groups to provide access to rapidly-changing information inaccessible or unavailable offline (26, 27) was characteristic of our participants’ experiences. For the majority of our participants, online Long Covid communities were a place to relate and empathise with others, similar to other communities whose illness experiences may have been medically contested (28, 29). Online communities have been described as pooling collective knowledge derived from the lay expertise of members with a vested interest in advancing the self-management of their condition (25). The information being shared is thus vetted and validated by the online community itself (30,31). In the Long Covid forums described by our participants, lived experience became more valuable than medical advice available offline. The credibility of “expert patients” (32) within online Long Covid forums might have been enhanced by the presence of many medical professionals living with Long Covid and acting as key contributors to these communities (33). Nevertheless, the novelty of Long Covid also meant that the lay expertise of members was sometimes dependent on learning from patients with other apparently similar conditions. In the case of graded exercise therapy, this had potential to cause confusion where advice was controversial or disputed by some patients.

### Limitations

This study aimed to provide in-depth exploration of the lived experience of Long Covid. Qualitative research of this kind necessitates a small sample size which naturally limits the generalisability of the research. We took steps to recruit a broad sample in terms of age, gender and ethnicity, but our participants were recruited from a research interest database indicating a level of engagement and access to online research that might not be representative of the Long Covid population as a whole. Future studies should seek to represent those from digitally excluded populations (34) in lived experience accounts of Long Covid, to further understand social and cultural sensitivities surrounding the experience.

## CONCLUSIONS

This study provides insight into the challenges of managing physical activity alongside the extended symptoms associated with Long Covid. Findings highlight the need for greater consensus around physical activity-related advice for people with Long Covid and improved support to resume activities considered important for wellbeing. The rapid and highly motivated ability of online communities to become trusted sources of information for self-management is also highlighted.

## Data Availability

Complete transcripts are not available as they pose a risk to participant confidentiality. All other study materials are available on reasonable request.

## Acknowledgments

The authors thank all participants of this study for sharing their time and experiences.

## Author statement

HH: developed the research question, participant recruitment, data collection, data analysis, manuscript preparation

LK and NK: data analysis; manuscript preparation

RC: developed the research question, secured funding for the research, acted as project advisor and manuscript review

## Notes

### Competing Interest Statement

The authors have declared no competing interest.

### Funding Statement

This work was supported by Sheffield Hallam University. No external funding was received.

### Author Declarations

Sheffield Hallam University Research Ethics Committee - reference ER25413313

